# Efficacy and safety of intensified versus standard prophylactic anticoagulation therapy in patients with Covid-19: a systematic review and meta-analysis

**DOI:** 10.1101/2022.03.05.22271947

**Authors:** Nicola K Wills, Nikhil Nair, Kashyap Patel, Omaike Sikder, Marguerite Adriaanse, John Eikelboom, Sean Wasserman

**Author notes:** **Corresponding author:** Nicola K Wills, Department of Medicine, Groote Schuur Hospital, University of Cape Town, Observatory, Cape Town, South Africa.

## Abstract

**Background:** Randomised controlled trials (RCTs) have reported inconsistent effects from intensified anticoagulation on clinical outcomes in Covid-19. We performed an aggregate data meta-analysis from available trials to quantify effect on non-fatal and fatal outcomes and identify subgroups who may benefit.

**Methods:** We searched multiple databases for RCTs comparing intensified (intermediate or therapeutic dose) versus standard prophylactic dose anticoagulation in adults with laboratory-confirmed Covid-19 through 19 January 2022. The primary efficacy outcome was all-cause mortality at end of follow-up or discharge. We used random effects meta-analysis to estimate pooled risk ratios for mortality, thrombotic, and bleeding events, and performed subgroup analysis for clinical setting and dose of intensified anticoagulation.

**Results:** Eleven RCTs were included (n = 5873). Intensified anticoagulation was not associated with a reduction in mortality for up to 45 days compared with prophylactic anticoagulation: 17.5% (501/2861) died in the intensified anticoagulation group and 18.8% (513/2734) died in the prophylactic anticoagulation group, relative risk (RR) 0.93; 95%CI, 0.79 – 1.10. On subgroup analysis, there was a possible signal of mortality reduction for inpatients admitted to general wards, although with low precision and high heterogeneity (5 studies; RR 0.84; 95% CI, 0.49 - 1.44; I^2^ = 75%) and not significantly different to studies performed in the ICU (interaction P = 0.51). Risk of venous thromboembolism was reduced with intensified anticoagulation compared with prophylaxis (8 studies; RR 0.53, 95%CI 0.41 – 0.69; I^2^ = 0%). This effect was driven by therapeutic rather than intermediate dosing on subgroup analysis (interaction P =0.04). Major bleeding was increased with use of intensified anticoagulation (RR 1.73, 95% CI 1.17 – 2.56) with no interaction for dosing and clinical setting.

**Conclusion:** Intensified anticoagulation has no effect on short term mortality among hospitalised adults with Covid-19 and is associated with increased risk of bleeding. The observed reduction in venous thromboembolism risk and trend towards reduced mortality in non-ICU hospitalised patients requires exploration in additional RCTs.

**Summary:** In this aggregate data meta-analysis, use of intensified anticoagulation had no effect on short term mortality among hospitalised adults with Covid-19 and was associated with increased risk of bleeding.

## Introduction

Covid-19 is associated with increased risk of venous and arterial thrombotic events,[1,2] particularly in patients with severe disease,[3] with incidence rates even higher than those seen in historical cohorts of critically-ill individuals with non-Covid-19 respiratory disease.[4] Venous thrombotic risk remains high even with use of standard prophylactic anticoagulation.[3] The interplay of direct viral-induced endothelial injury with a dysregulated inflammation response and coagulation factor activation are postulated as key contributors to the development of the Covid-19-associated prothrombotic state.[5–7] Thrombo-inflammation has been linked to disease progression and poor outcomes in patients with Covid-19;[6,8] in particular, increased circulating D-dimer (a biomarker of inflammation and coagulation activation) is an independent predictor of mortality.[9–11]

These observations led to widespread use of therapeutic anticoagulation in patients hospitalized with Covid-19, especially heparin, which is believed to have anti-inflammatory and anti-viral properties,[12,13] in the hope it may prevent thrombotic events and improve outcomes. Some non-comparative studies suggested that intensified (intermediate or therapeutic) dose anticoagulation may reduce thrombotic complications,[14,15] but cohort studies with matched controls did not show mortality benefit[16,17] and higher bleeding risk has been consistently reported.[18,19] Observational studies are limited by the potential for confounding as well as non-comparability across study populations, selection and observer bias, and inconsistent ascertainment of key outcomes, leaving major uncertainty around risk-benefit.

Randomized controlled trials (RCTs) offer more robust estimates of treatment effect. However, most RCTs of anticoagulation strategies for Covid-19 have been small, enrolling several hundred rather than thousands of participants, and were not powered to assess important individual clinical outcomes. Three RCTs, enrolling between three and seven hundred participants per treatment arm, were neutral for primary composite outcomes that included both thrombotic events and mortality, and did not demonstrate mortality benefit with intensified anticoagulation, and only one of these trials showed a reduction in thrombotic events.[20–22] A larger RCT involving non-critically ill patients (n = 2,219) hospitalized with Covid-19 found that intensified therapy compared with usual dose thromboprophylaxis reduced need for organ support and major thrombosis, but not overall mortality. A small effect with low precision in this single positive trial, inconsistent effects across different studies, and a strong reproducible signal of increased bleeding risk limits definitive conclusions around use of intensified anticoagulation in Covid-19.[23] Synthesizing evidence from all available RCTs may provide more precise estimates of effect and identify subgroups that derive the greatest absolute benefit from intensified anticoagulation. Additional power from pooled data may also enable separate examination of the effects of treatment on individual outcomes, for example, thrombotic events and mortality, potentially providing insights into the prognostic importance of thrombosis. We undertook a systematic review and aggregate data meta-analysis to obtain best estimates of the effect of intensified versus standard prophylactic anticoagulation on clinically important outcomes for patients with Covid-19.

## Methods

### Eligibility criteria

We included RCTs comparing intensified, defined as intermediate (generally 1 mg/kg of enoxaparin once daily, or an equivalent) or therapeutic dosing, versus standard prophylactic dose anticoagulation for adults with laboratory-confirmed Covid-19 (Table 1). No restriction by language, publication status (including articles in pre-print), anticoagulation agent, or clinical setting was applied (supplementary Table S1). We only included studies reporting at least one of the prespecified outcomes listed in Table 1.

**Table 1.**
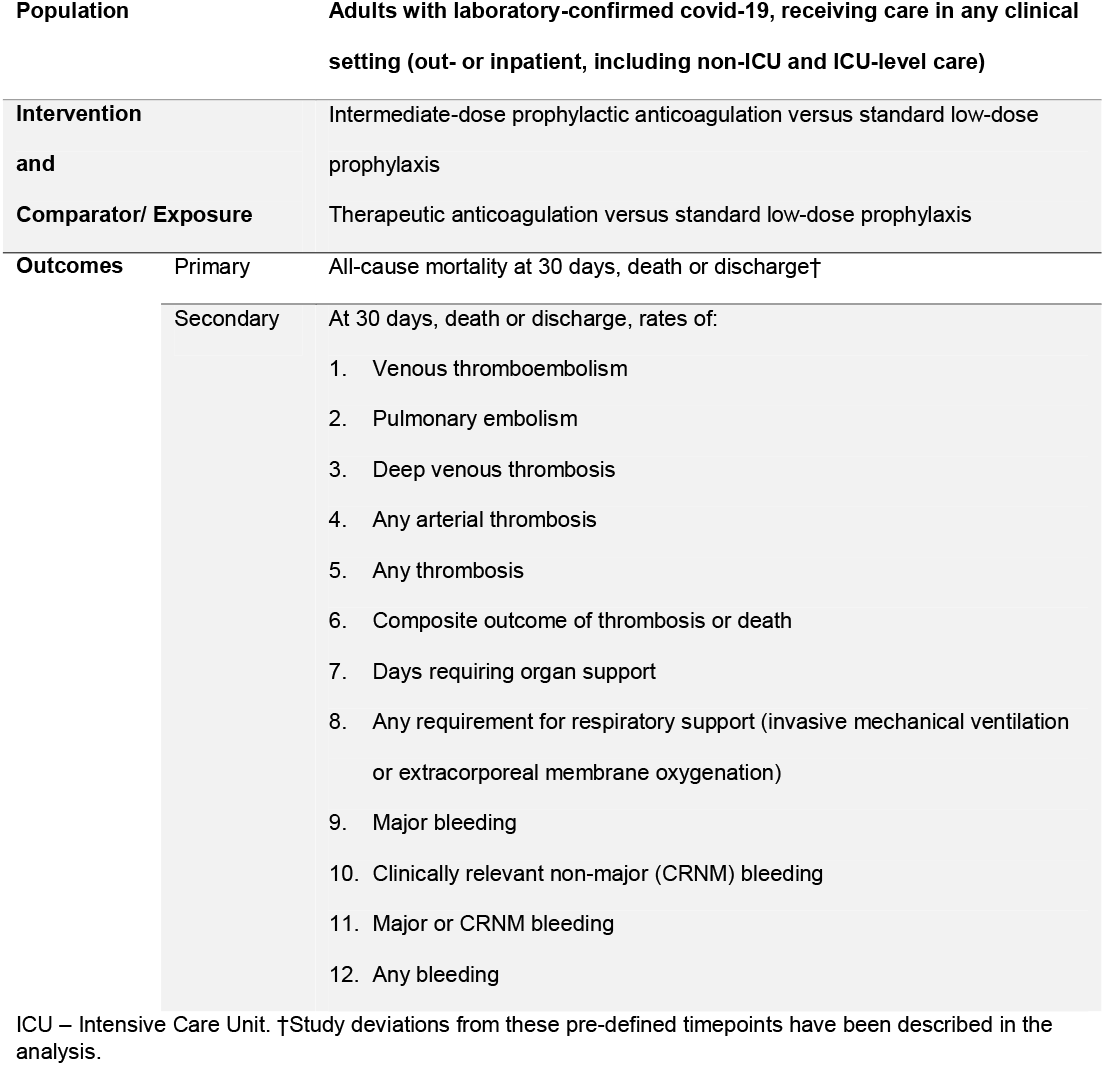
PICOT Eligibility criteria for study inclusion.

### Search strategy

An electronic search was conducted on 19 September 2021 and repeated on 19 January 2022 using MEDLINE (PubMed), Scopus, the World Health Organization (WHO) COVID-19 database (https://search.bvsalud.org/global-literature-on-novel-coronavirus-2019-ncov/), and the Cochrane Library. We also screened the WHO Trial Registry Network (https://trialsearch.who.int/) and ClinicalTrials.gov (https://clinicaltrials.gov/) for ongoing/recently completed trials, and PROSPERO (https://www.crd.york.ac.uk/PROSPERO/) for ongoing or recently completed systematic reviews. We searched preprint literature by scanning the WHO COVID-19 database as well as the National Institutes of Health *iSearch* COVID-19 portfolio (https://icite.od.nih.gov/covid19/search/). A search strategy was developed using multiple terms relating to anticoagulation, anticoagulant agents, and Covid-19 (supplementary Table S2).

### Record management and data extraction

Records from the primary search were entered into Mendeley Reference Management Software Version 1.19.8 (https://www.mendeley.com/) and duplicates removed. Titles and abstracts were screened against the study eligibility criteria (Table 1) by KP, NH, and OS and independently by MA and NW, followed by review of the full texts of potentially eligible articles for inclusion. After consensus on studies meeting criteria for inclusion, variables of interest (supplementary Table S3) were extracted on a Microsoft Excel spreadsheet by NH and OS with independent verification by MA and NW. Reference lists of included studies were screened to identify any additional eligible studies. Risk of bias in individual studies was independently assessed by KP, MA and NW using version 2 of the Cochrane risk-of-bias tool for randomized trials (https://training.cochrane.org/handbook/current/chapter-08), with respect to the key outcome of interest (mortality). SW/JE were consulted for review of any conflict regarding study inclusion, data discrepancies, or assessing risk of bias.

### Data analysis

The primary outcome was all-cause mortality at end of follow-up or discharge. Other efficacy outcomes of interest included venous thromboembolism (symptomatic or asymptomatic VTE, including pulmonary embolism (PE) or deep vein thrombosis (DVT)), arterial thrombosis (stroke, myocardial infarction, acute limb ischemia, other arterial ischemia), any thrombotic event, and a composite of thrombosis or death. The key safety outcome was major bleeding; other safety outcomes included clinically relevant non-major bleeding and any bleeding event. We planned to analyse the effect of intensified anticoagulation on days requiring any organ support and respiratory support (invasive mechanical ventilation or extracorporeal membrane oxygenation) but these outcomes were not reported by included trials.

We performed an intention-to-treat analysis (the denominator was all randomized participants who received at least one dose of assigned treatment). Data was pooled using a random effects meta-analysis model with restricted maximum likelihood estimation. We computed risk ratios (RR) with 95% confidence interval (95%CI) as measures of effect. Between-study heterogeneity was quantified using the I^2^ statistic.[24] Sensitivity analysis using the “leave-one-out” approach was done to visually evaluate the influence of each study on the overall pooled effect for mortality. We performed pre-specified subgroup analysis for baseline severity of illness (intensive care unit (ICU) setting versus general ward [where >50% of randomised participants admitted in general ward]) and dose of intensified anticoagulation (therapeutic versus intermediate doses). Funnel plots were generated to assess publication bias for each of the primary and secondary outcomes. All meta-analyses were performed using Stata 17.

This study is registered on PROSPERO (ref <u>CRD42021273449</u>).

## Results

### Characteristics of included studies

We screened 2470 records and included 11 studies meeting eligibility criteria (Figure 1); these studies contributed data from 5873 adults with confirmed Covid-19 who were followed up over a median of 30 days (range 21 to 45 days). Key information from included studies is summarised in Table 2 with full study details provided in Supplementary Tables S4 – S6.

**Figure 1.**
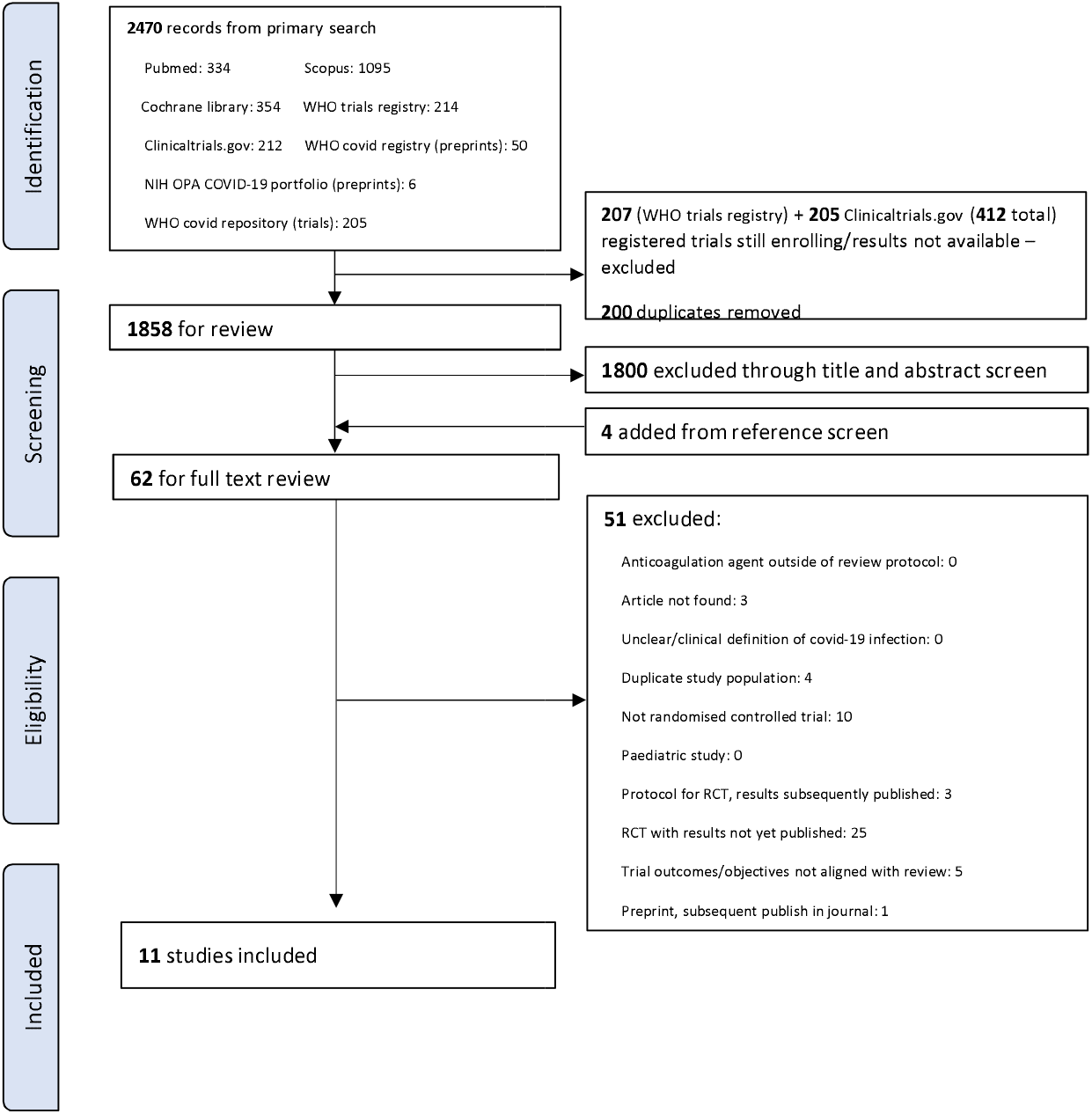
PRISMA diagram.

**Table 2.**
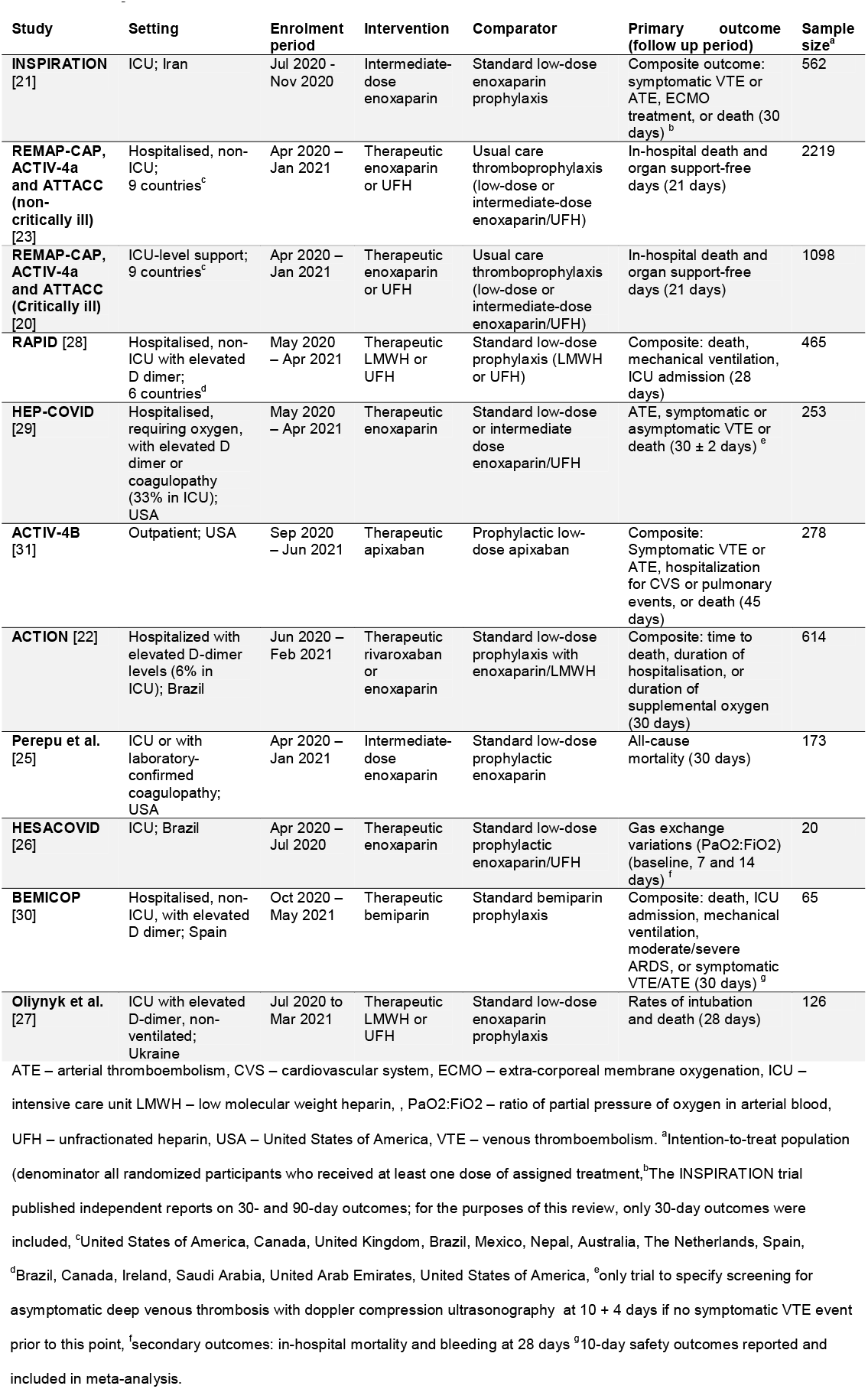
Key details of included studies.

Five ICU-based studies reported outcomes among 1979 critically-ill patients, [20,21,25–27] five studies reported outcomes from 3616 patients hospitalised in a general ward setting, [22,23,28–30] and one study reported outcomes from 278 outpatients.[31] Nine studies (n = 5138) [20,22,23,26–31] compared therapeutic low molecular weight heparin (LMWH), unfractionated heparin (UFN), or rivaroxaban/apixaban to standard thromboprophylaxis (three inpatient studies allowed either standard low-dose or intermediate-dose enoxaparin in the ‘usual care’ comparator arm [20,23,29]). In the remaining two studies (n = 735),[21,25] both conducted in an ICU setting, intermediate dose enoxaparin was compared to standard dose enoxaparin thromboprophylaxis.

Median age range was 52 to 71 years and 41% were female (11 studies, n = 5873) with median BMI 26 to 34 kg/m^2^ (10 studies, n = 5747). 38% were prescribed an antiviral agent at baseline (8 studies, n = 5004) and 64% received corticosteroids at baseline (9 studies, n = 5469). Hypertension was reported in 45% (9 studies, n = 4659) and diabetes in 30% (10 studies, n = 5747). Chronic lung or cardiovascular disease were documented in 17% and 8%, respectively (9 studies, n = 5469).

Risk of bias assessment is reported in the Supplementary Material (Table S7 and Figure S1): 4 studies had a low risk of bias, 2 were assessed as high risk, and 5 had some concerns. Funnel plot for the mortality outcome showed some asymmetry, suggesting possible publication bias, but the number of included studies was small (Supplementary Figures S2)

### Primary outcome

Eleven studies were included for the primary outcome of all-cause mortality: 16.7% (501/3004) died in the intensified anticoagulation group and 17.9% (513/2869) died in the prophylactic anticoagulation group. Intensified anticoagulation was not associated with a reduction in mortality for up to 45 days compared with prophylactic anticoagulation (RR 0.93; 95%CI, 0.79 – 1.10). There was significant heterogeneity, with 37% of variability in effect size estimates due to between-study differences (P = 0.03) (Figure 2a). On sensitivity analysis omission of individual trials had no significant influence on pooled mortality (Supplementary Figure S3).

**Figure 2a.**
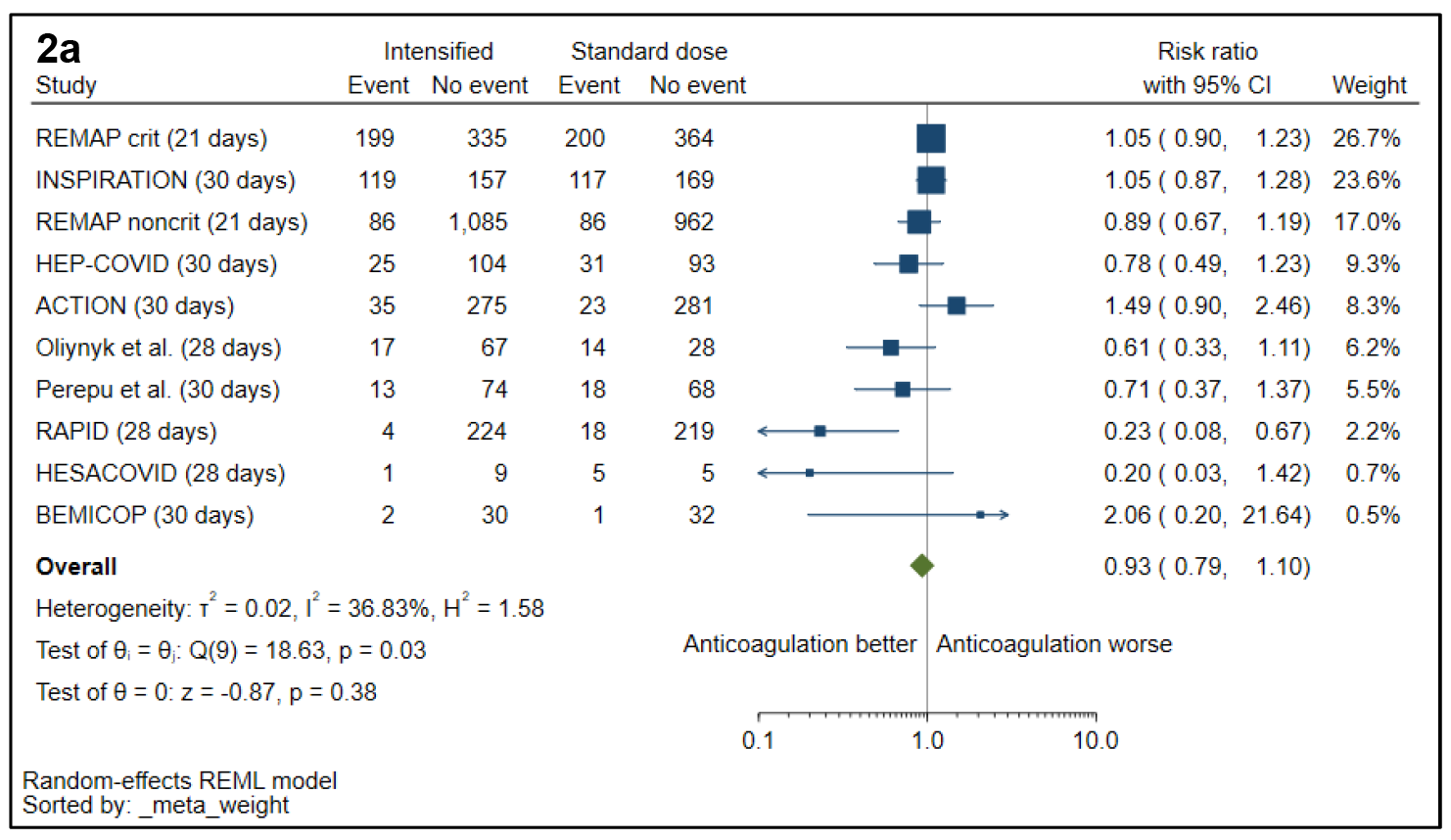
Mortality with intensified versus prophylactic anticoagulation. The single outpatient trial [31] was excluded from the forest plot because of no mortality events.

### Secondary efficacy outcomes

Only one study (n = 253)[29] screened for asymptomatic DVT with doppler compression ultrasonography, but the majority of reported VTE events were symptomatic. Symptomatology was not specified in the REMAP-CAP platform of two multi-centre trials.[20,23] The remaining studies reported rates of symptomatic VTE (n = 4207) (see Table 2). Risk of VTE was consistently reduced with intensified anticoagulation compared with prophylaxis: 2.8% [81/2888] versus 5.4% [151/2794]; RR 0.53, 95%CI 0.41 – 0.69; I^2^ = 0%, 8 studies) (Figure 2b). The effect was driven by a reduction in PE (1.3% [37/2801] vs 3.5% [95/2708]; RR 0.39, 95%CI 0.27 – 0.57; I^2^ = 0%) but not DVT (1.3% [36/2801] vs 1.7% [47/2708]; RR 0.81, 95%CI 0.48 – 1.35; I^2^ = 21%) (Supplementary Figure S4 – S5). Intensified anticoagulation was also associated with a reduction in the composite outcome of thrombosis or death (4 studies, RR 0.78; 95% CI, 0.66 – 0.91; I^2^ = 0%) (Supplementary Figure S6). Risk for any thrombosis was reduced (Supplementary figure S7), but without evidence of effect on arterial thrombosis (8 studies, RR 1.26; 95% CI, 0.57 - 2.77; I^2^ = 50%).

**Figure 2b.**
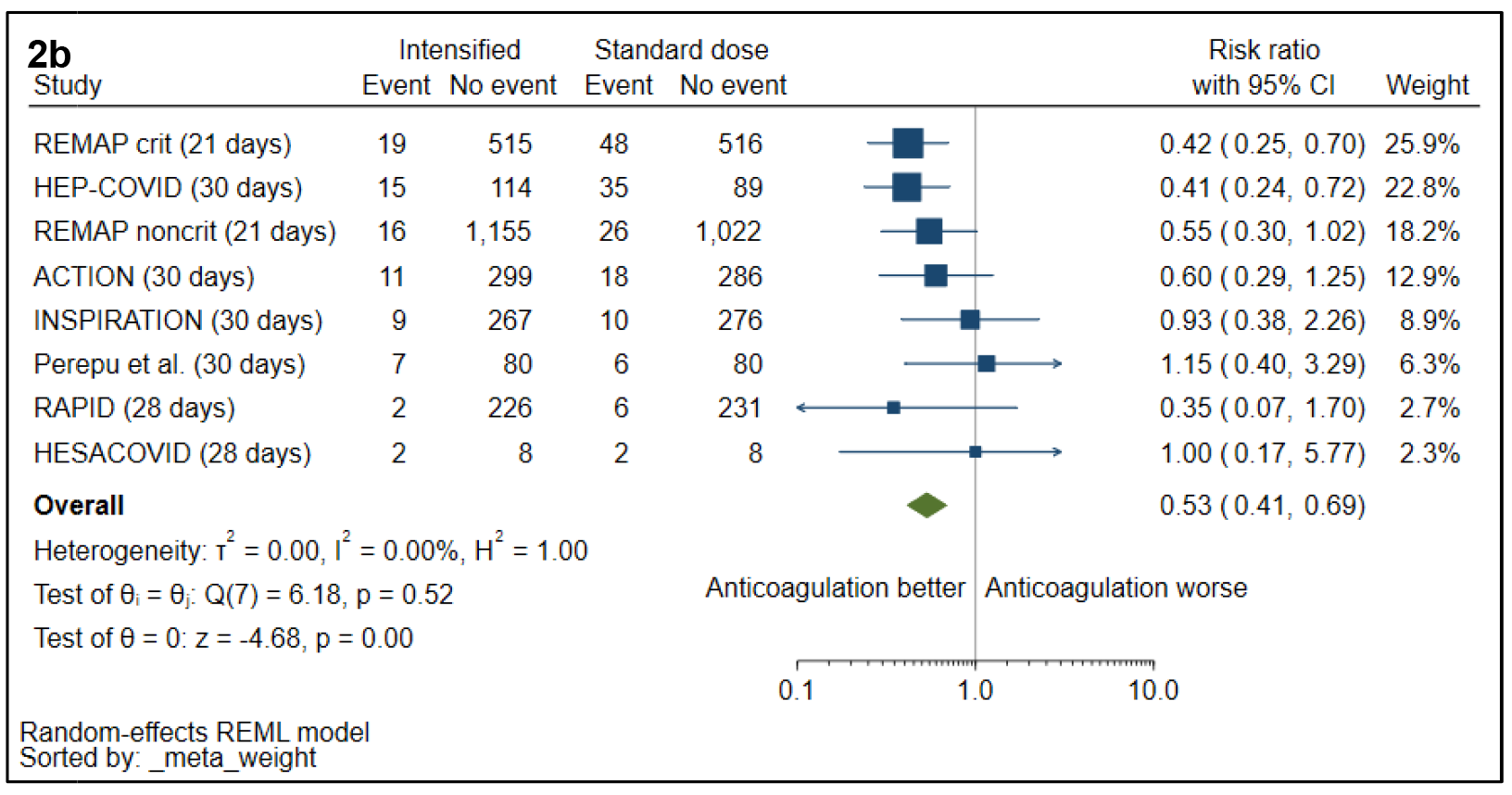
Venous thromboembolism with intensified versus prophylactic anticoagulation. The single outpatient trial [1] was excluded from the forest plot because of no mortality events. Two other trials were excluded because venous thromboembolic events were not captured as outcomes [27,30].

### Safety outcomes

Risk of major bleeding was increased with intensified anticoagulation compared with prophylaxis: 2.3% [69/3004] vs 1.3% [38/2869]; RR 1.73, 95% CI 1.17 – 2.56, I^2^ = 0%; 11 studies) (Figure 2c). Risk of clinically relevant non-major bleeding (4.4% vs 1.9%; 7 studies; RR 2.08, 95% CI 1.13 – 3.83; I^2^ = 11%) and any bleeding (8.8% vs 4.3%; 7 studies; RR 1.90, 95% CI 1.16- 3.12; I^2^ = 30%) was also increased with use of intensified anticoagulation (Supplementary Figures S7 – S11)

**Figure 2c.**
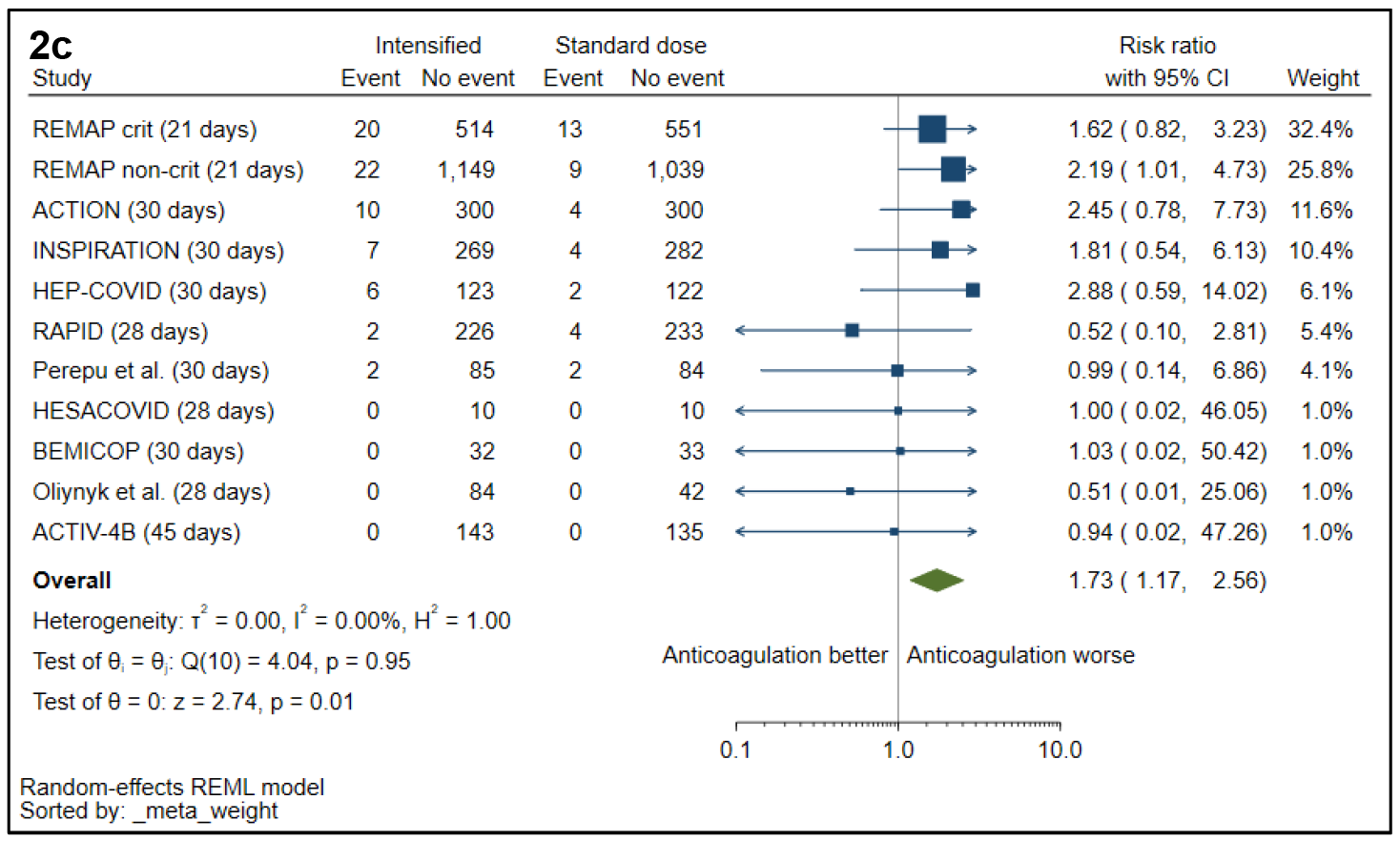
Major bleeding with intensified versus prophylactic anticoagulation.

### Subgroup analysis

There was a signal of mortality reduction for inpatients admitted to general wards, although with low precision and high heterogeneity (5 studies; RR 0.84; 95% CI, 0.49 - 1.44; I^2^ = 75%); this effect was not significantly different to studies performed in the ICU (interaction P = 0.51) (Figure 3a). There was also no difference in effect between therapeutic and intermediate dosing on mortality (interaction P = 0.46), but substantial heterogeneity existed between trials testing therapeutic doses (I^2^ = 67%, P = 0.02) (Figure 3b).

**Figure 3.**
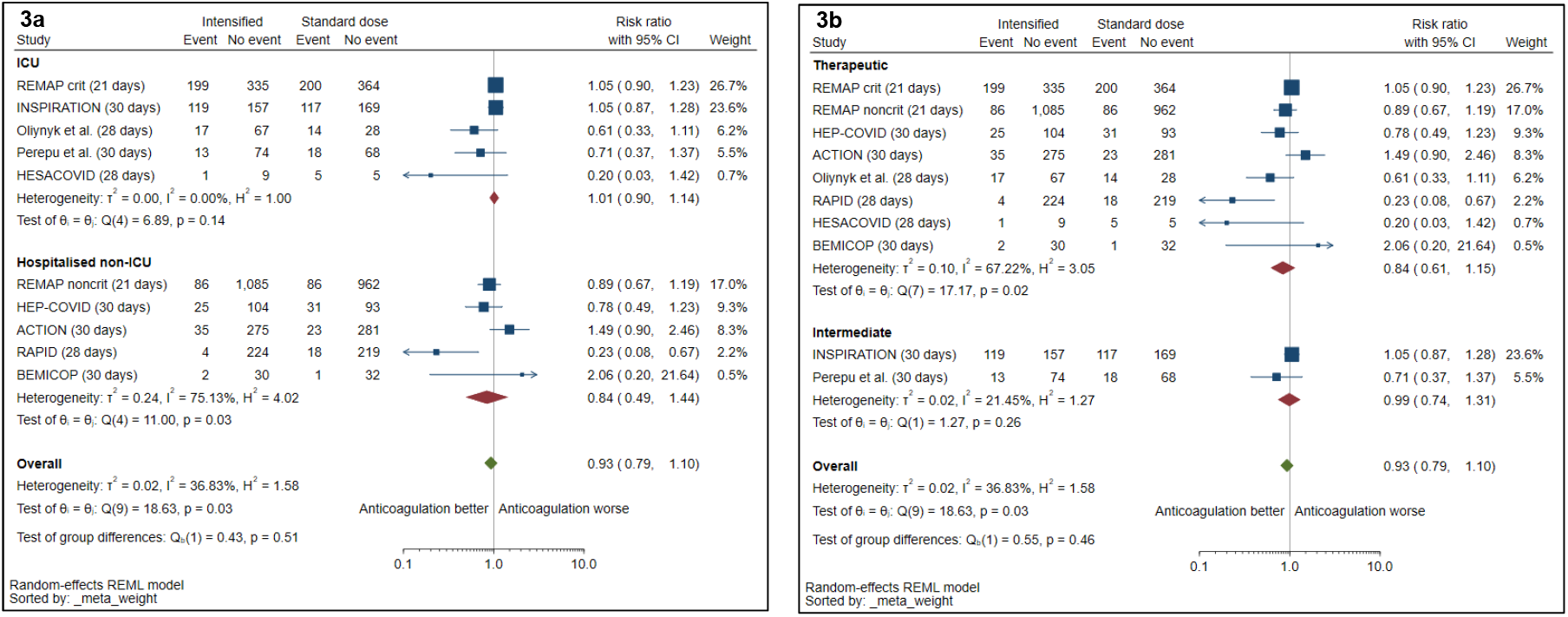
Subgroup analysis of mortality with intensified versus prophylactic anticoagulation. (a) by clinical setting (ICU versus hospitalised non-ICU) and (b) by dose of intensified anticoagulation (therapeutic versus intermediate). The single outpatient trial [31] was excluded from the forest plot because of no mortality events. Two other trials were excluded because venous thromboembolic events were not captured as outcomes [27,30].

Pooled VTE risk reduction was greater in studies conducted in hospitalised non-ICU settings (4 studies; RR 0.49, 95%CI 0.34 – 0.69) compared with those done in ICU (4 studies; RR 0.70, 95% CI 0.38 – 1.28), but this difference was not statistically significant (interaction P = 0.31) (Figure 4). This effect was seen in trials using therapeutic anticoagulation (6 studies; RR 0.47, 95% CI 0.36 – 0.63) but not those testing intermediate-dose anticoagulation (2 studies; RR 1.02, 95% CI 0.52 – 2.0); interaction P = 0.04 (Supplementary Figure S12). In an exploratory analysis, there was no reduction in mortality with intensified anticoagulation in both trials showing a significant reduction in VTE events among non-critically ill patients (REMAP-CAP/ACTIV-4a/ATTACC non-critically ill and HEP-COVID, n = 2,472; RR 0.86; 95% CI, 0.67 – 1.10; I^2^ = 0%) or in trials without a clear VTE effect (RR 0.62; 95% CI, 0.10 – 3.87; I^2^ = 90%).

**Figure 4.**
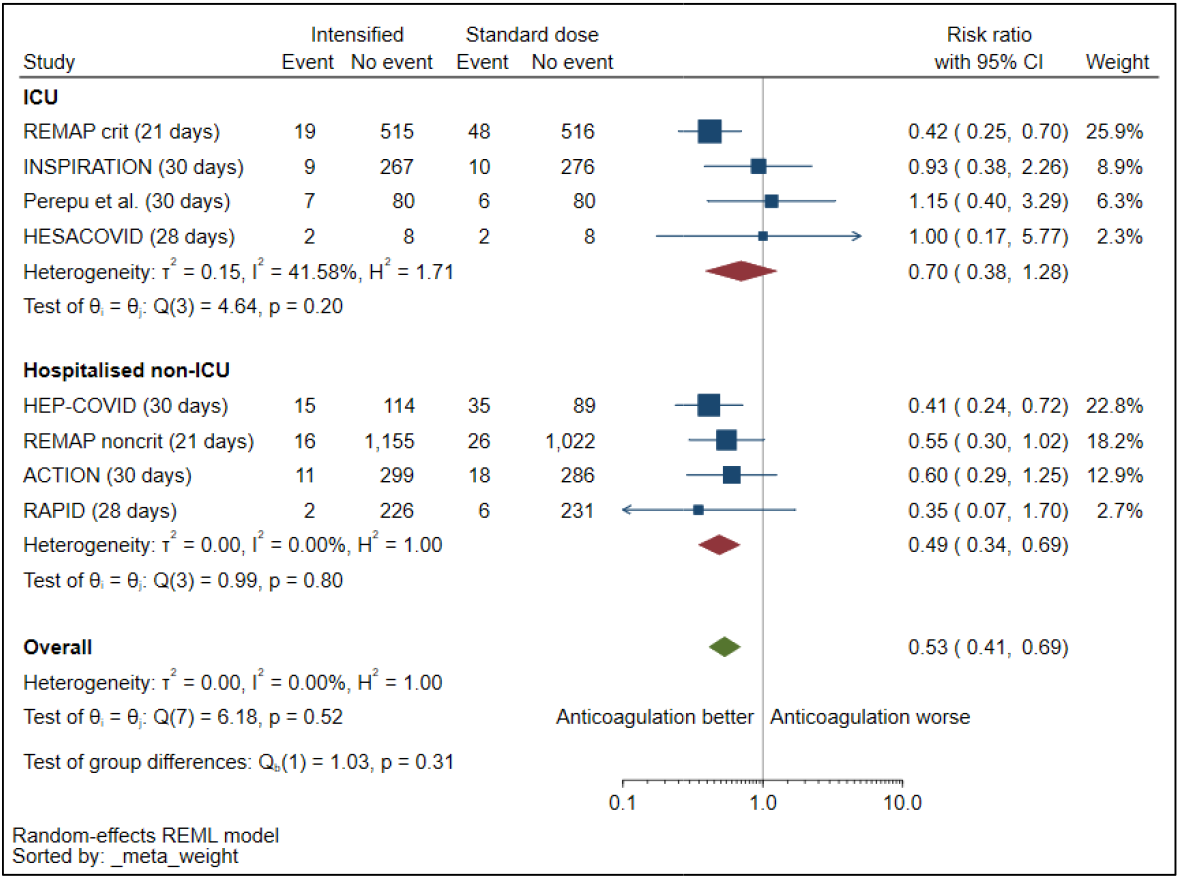
Subgroup analysis of VTE with intensified versus prophylactic anticoagulation, by stratified by clinical setting (ICU versus hospitalised non-ICU). There were no reported thrombotic events in the single outpatient trial[31] and venous thromboembolic events were captured as outcomes in the remaining two trials[27,30] – these trials are excluded from the forest plot above.

Similar increases in major bleeding were observed among critically ill and non-critically ill patients (interaction P = 0.55) and those receiving therapeutic versus intermediate anticoagulant dosing (interaction P = 0.80) (Figures 5a and 5b).

**Figure 5.**
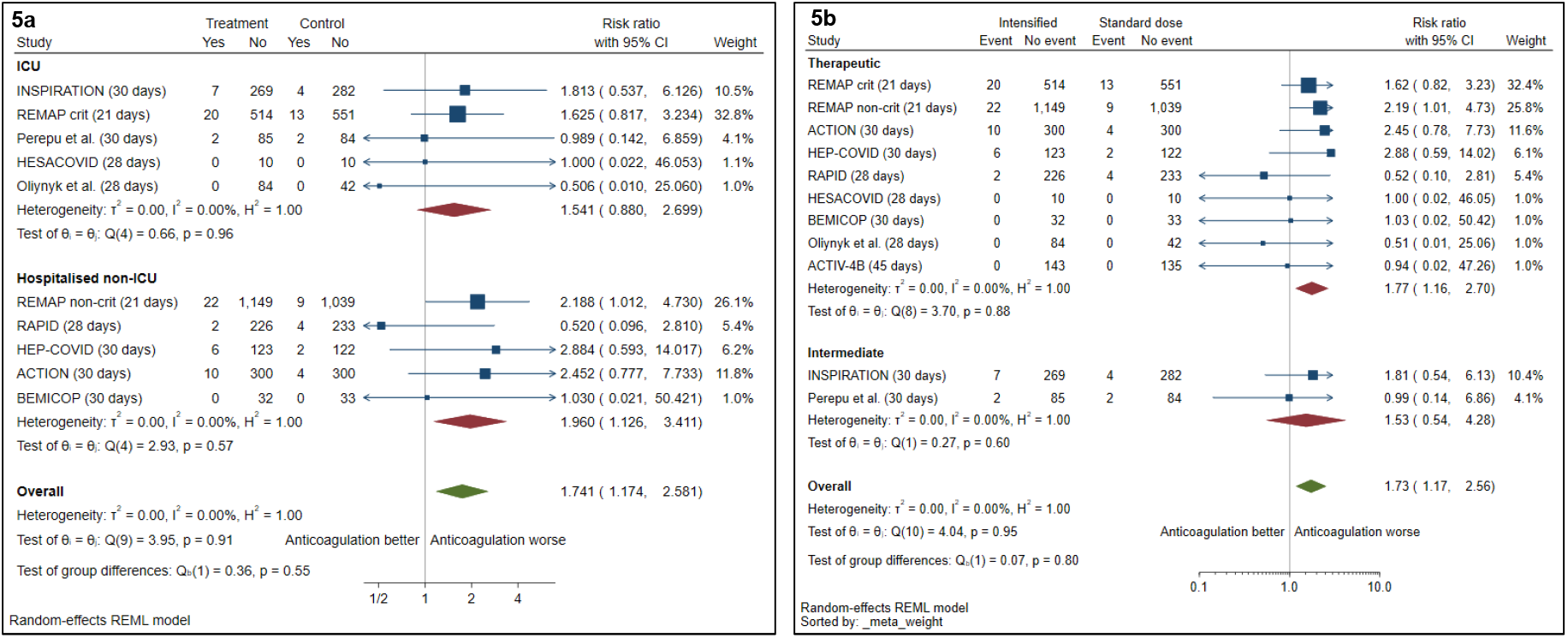
Subgroup analysis of major bleed with intensified versus prophylactic anticoagulation. (a) by clinical setting (ICU versus hospitalised non-ICU) and (b) by dose of intensified anticoagulation (therapeutic versus intermediate).

## Discussion

The data from this meta-analysis, synthesizing outcomes from 11 RCTs involving 5873 adults, show that intensified anticoagulation did not reduce short term mortality (up to 45 day) for hospitalised patients with Covid-19. This finding was consistent across the spectrum of clinical severity and anticoagulant dosing strategies. Intensified anticoagulation reduced VTE as well as the composite outcome of VTE and death, but at a cost of significantly increased risk of major bleeding.

Covid-19 pneumonia is associated with a hypercoagulable state resulting from endothelial perturbation and an intense prothrombotic inflammatory response.[32] This may progress to a distinct syndrome, termed Covid-19-associated coagulopathy, characterised by markedly elevated D-dimer and fibrinogen concentrations and pulmonary microvascular thrombosis, which has been linked with worse outcome.[5,7–11,33,34] VTE is common even with use of standard dose thromboprophylaxis, possibly occurring at higher rates than other respiratory conditions.[1] Given the prominence of thrombo-inflammation in the pathogenesis of Covid-19 and the likelihood that pulmonary thrombotic complications contribute to progressive hypoxic respiratory failure, one might expect that by preventing VTE, intensified dosing of anticoagulation should reduce disease severity and related mortality. The lack of overall survival benefit despite significant reduction in VTE events with intensified anticoagulation observed across high quality trials in our meta-analysis therefore requires explanation.

Our findings are consistent with evidence from medical inpatients without Covid-19, where thromboprophylaxis has established benefit for preventing VTE regardless of risk and illness severity,[35–37] but does not reduce mortality and its effect on other important clinical outcomes, such as symptomatic PE, is uncertain.[38] Several factors could play a role in this apparent paradox. Most trials of anticoagulation, including for Covid-19, are not powered to detect a difference in mortality, and absence of an effect on this outcome may result from type 2 error rather than true lack of efficacy. Related to this, thrombotic events, often ascertained as venographic DVT with uncertain clinical significance, are inadequate as a surrogate for efficacy outcomes in thromboprophylaxis trials because of poor correlation with important outcomes[39] - although prophylaxis prevents thrombotic events overall, trials may fail to detect an effect on fatal PE.

There are plausible biological explanations for true absence of mortality effect. The increased risk of major bleeding associated with thromboprophylaxis - 80% for standard heparin doses in the most recent Cochrane review [38] and an additional 74% increased risk from intensified anticoagulation for Covid-19 in our analysis - may offset any reduction in mortality due to VTE. Although risk of overt bleeding from intensified anticoagulation was increased in both non-ICU and ICU settings, alveolar haemorrhage, which has been documented in Covid-19-associated ARDS, [40] may also contribute to overall harm, especially in the latter group. Another possibility is that intensified prophylaxis, even at therapeutic doses, may not lead to reduction in fatal PE and translate into mortality benefit. This is especially relevant in ICU settings where a larger proportion of non-VTE-attributable deaths occur and the presence of ARDS-associated pulmonary microvascular thrombosis (‘immunothrombosis’) may be refractory to heparin therapy. Although intensified anticoagulation does reduce PE events this may not an important cause of death in Covid-19, limiting impact on mortality.

An advantage of meta-analysis is the potential to identify subgroups not observed in individual trials that may benefit from an intervention. Our analysis found significant reductions in VTE only in trials that included non-critically ill patients (which all provided therapeutic doses of anticoagulation); this was accompanied by a signal of mortality reduction not seen in trials conducted in the ICU, although with significant between-study heterogeneity. Smaller meta-analyses investigating anticoagulation in Covid-19 have also reported a trend towards reduced mortality in non-critically ill patients only [41–43]. These findings suggest that a window may exist earlier in the disease course of Covid-19 for optimal timing of anticoagulation to prevent VTE and avert disease progression via reduction of pulmonary microthrombosis and pleotropic effects of heparin. The average number of days from symptom onset to hospitalisation or enrolment ranged from 1.4 to 10 days among included studies in our review, and 4 of the 5 trials in non-ICU settings required elevated D-dimer or other indicator of coagulopathy for enrolment. These patients may have already developed Covid-19-associated coagulopathy, possibly missing a crucial intervention period where benefit of anticoagulation may be maximised. Currently, however, the absence of demonstrable effect on mortality coupled with significantly increased bleeding risk (which includes intracranial and fatal bleeding in some trials) does not justify introduction of intensified anticoagulation into routine care for non-critically ill patients with Covid-19 pneumonia.

Existing data also do not provide clear guidance for an optimal anticoagulation dosing strategy that balances risk of bleeding with clinical benefit. On subgroup analysis, the largest effect on VTE reduction (Fig. S12) was seen with therapeutic doses of anticoagulation. Bleeding risk was statistically similar across dosing groups, but the precision was low for intermediate dosing and the established dose-response relationship for bleeding with heparin raises concerns about use of therapeutic dosing. There is currently no RCT data on use of intermediate-dose anticoagulation for Covid-19 in non-critically ill adults, who appeared to derive the most benefit from anticoagulation. Although VTE reduction was only apparent in trials using therapeutic anticoagulation, observational studies have suggested mortality benefit and lower bleeding risk from intermediate-dose anticoagulation among hospitalised Covid-19 patients, with a high representation of patients from general wards.[44,45] Ongoing trials predominantly enrolling non-critically ill adults will inform the role and optimal use of intensified prophylaxis in Covid-19: ASCOT (<u>NCT04483960</u>, n = 2400, therapeutic and intermediate LMWH versus standard prophylaxis); PROTHROMCOVID (<u>NCT04730856</u>, n = 600, therapeutic and intermediate tinzaparin versus standard prophylaxis); INHIXACOV19 (<u>NCT04427098</u>, n = 300, intermediate versus prophylactic dose enoxaparin); XACT (<u>NCT04640181</u>, n = 150, therapeutic or intermediate enoxaparin or rivaroxaban versus standard prophylaxis); ACT (<u>NCT04324463</u>, n = 6000, aspirin and rivoraxaban versus standard of care); and FREEDOM COVID (<u>NCT04512079</u>, n = 3600, therapeutic enoxaparin versus enhanced dose rivaroxaban versus prophylaxis).

This review has several limitations. First, we analysed trial-level data, limiting the extent to which we could explore differences in subgroups by important baseline prognostic variables such as age, comorbidity, and markers of disease severity and inflammation. Second, although we performed subgroup analysis by clinical setting (as a surrogate for disease severity), criteria for severe disease and ICU eligibility were institution and study-specific, limiting generalisability. This may have contributed to the extreme heterogeneity (I^2^ = 75%) observed among non-ICU-based studies in the risk ratios for mortality. Third, the relatively small number of events limited precision of effect estimates, especially for the non-critically ill subgroup where there was possibly a signal for reduced mortality. We were not able to analyse effect of intensified anticoagulation on need for, and duration of, organ support since these outcomes were not consistently reported. Fourth, we identified 2 studies to be at high risk of bias and with some concerns, chiefly with regards trials using non-objective methods in defining and detecting thrombosis events. This serves to emphasise the limitation using of thrombotic events as an outcome in anticoagulation trials. Fifth, asymmetry in the funnel plots indicates possibility of publication bias, but the small number of included trials limits accuracy. Finally, although sensitivity analysis showed no effect modification on the primary outcome with omission of individual trials, this meta-analysis was dominated by events from two large multi-centre studies [23][20] in which a large proportion of patients in the usual care groups received intermediate-dose prophylaxis. This may have skewed the effect of intensified anticoagulation toward the null; one recent systematic review showed a more precise effect of anticoagulation on mortality (albeit still non-significant) among moderately ill patients after excluding these trials.[28]

In conclusion, available data indicate that intensified anticoagulation has no effect on short term mortality among hospitalised adults with Covid-19 and is associated with increased risk of bleeding. The finding of significant reductions in VTE with a possible signal for reduced mortality in non-ICU hospitalised adults suggests additional studies, with a focus on moderately-ill patients and different dosing strategies, may delineate optimal use of thromboprophylaxis in this condition.

## Data Availability

This is a meta-analysis of data from randomised controlled trials with publicly available results (all available online with URLs as listed in reference lists)

## Acknowledgements

The authors would like to thank the University of Cape Town Health Sciences Library for assistance with development of search terms and strategy,

## Author contributions

Conception and writing of protocol: NW, SW, JE. Registration of protocol on PROSPERO: NH, KP, OS. Record screening, data extraction, and Risk of Bias assessment: NW, NH, KP. OS and MA. Analysis and interpretation: NW, SW, JE. Drafting of manuscript: NW, SW, JE. Critical review of the manuscript: all authors.

## Figure Legends

**Figure 1. PRISMA diagram**

**Figure 2a. Mortality with intensified versus prophylactic anticoagulation**.

The single outpatient trial [31] was excluded from the forest plot because of no mortality events.

**Figure 2b. Venous thromboembolism with intensified versus prophylactic anticoagulation**.

The single outpatient trial [31] was excluded from the forest plot because of no mortality events. Two other trials were excluded because venous thromboembolic events were not captured as outcomes [27,30].

**Figure 2c. Major bleeding with intensified versus prophylactic anticoagulation**.

**Figure 3. Subgroup analysis of mortality with intensified versus prophylactic anticoagulation**

(a) by clinical setting (ICU versus hospitalised non-ICU) and (b) by dose of intensified anticoagulation (therapeutic versus intermediate). The single outpatient trial [31] was excluded from the forest plot because of no mortality events. Two other trials were excluded because venous thromboembolic events were not captured as outcomes [27,30].

**Figure 4. Venous thrombosis with intensified versus prophylactic anticoagulation, by stratified by clinical setting (ICU versus hospitalised non-ICU)**.

The single outpatient trial [31] was excluded from the forest plot because of no mortality events.

Two other trials were excluded because venous thromboembolic events were not captured as outcomes [27,30].

**Figure 5. Subgroup analysis of major bleeding with intensified versus prophylactic anticoagulation**,

(a) by clinical setting (ICU versus hospitalised non-ICU) and (b) by dose of intensified anticoagulation (therapeutic versus intermediate).

