## Supplementary material for "Efficacy and safety of intensified versus standard prophylactic anticoagulation therapy in patients with Covid-19: a systematic review and meta-analysis"

**Supplementary Table S1. Definition of interventions**

| Therapeutic anticoagulation | Agents: unfractionated heparin (UFH), low molecular weight heparin (LMWH), fondaparinux, heparinoids (including sulodexide and dociparstat and nafamostat), parenteral direct thrombin inhibitors (DTIs) and direct oral anticoagulants (DOACs).  Dosing as per trial protocol, in alignment with international recommendations for the treatment of acute venous thromboembolism. |
| --- | --- |
| Intermediate-dose anticoagulation | Agents: unfractionated heparin (UFH), low molecular weight heparin (LMWH), fondaparinux, heparinoids (including sulodexide and dociparstat and nafamostat), parenteral direct thrombin inhibitors (DTIs) and direct oral anticoagulants (DOACs).  Dosing as per study or local protocol, at doses higher than that required for prevention of acute venous thromboembolism, but not reaching dosing required for treatment of confirmed venous thromboembolism as per international guidelines. |
| Standard low-dose prophylaxis | Agents: unfractionated heparin (UFH), low molecular weight heparin (LMWH), fondaparinux, heparinoids (including sulodexide and dociparstat and nafamostat), parenteral direct thrombin inhibitors (DTIs) and direct oral anticoagulants (DOACs).  Dosing as per trial protocol, in alignment with international recommendations for the prevention of acute venous thromboembolism. |
| Intensified anticoagulation | Therapeutic or intermediate-dose anticoagulation |
| Organ support | Non-invasive (including high flow nasal cannula support), invasive mechanical ventilation, extracorporeal membrane oxygenation, inotrope or vasopressor support, renal replacement therapy).  ICU-level support including care offered within ICU as well as organ-support offered in non-ICU settings. |

**Supplementary Table S2. Search terms, as used in Pubmed and adapted for use on other platforms**

| Search query | Terms |
| --- | --- |
| #1 | Anticoagulation OR anticoagulant OR Thromboprophylaxis OR Antithrombotic OR Anti-thrombosis |
| #2 | heparin OR UFH OR “unfractionated-heparin” OR LMWH OR “low molecular weight heparin” OR dalteparin OR enoxaparin OR nadroparin OR tinzaparin OR bemiparin OR certoparin OR parnaparin OR reviparin OR “indirect factor Xa inhibitor” OR fondaparinux OR heparinoids OR sulodexide OR dociparstat OR nafamostat OR NOAC OR DOAC OR “direct oral anticoagulant” OR “novel oral anticoagulant” OR “non-vitamin K antagonist oral anticoagulant” OR “direct factor Xa inhibitor” OR apixaban OR rivaroxaban OR edoxaban OR betrixaban OR “parenteral direct thrombin inhibitor” OR DTI OR “direct thrombin inhibitor” OR dabigatran OR bivalirudin OR argatroban OR “target-specific oral anticoagulant” OR TSOAC OR “oral direct inhibitor” OR ODI |
| #3 | Betacoronavirus OR “Corona Virus” OR “Corona Viruses” OR Coronavirus OR COVID OR “coronavirus disease-2019” OR “coronavirus 2019” OR COVID19 OR covid-19 OR “2019 novel coronavirus infection” OR “2019 novel coronavirus disease” OR “2019-nCoV disease”  OR CoV OR CoV2 OR HCoV-19 OR nCoV OR 2019nCoV  OR “severe acute respiratory syndrome CoV” OR “severe acute respiratory syndrome coronavirus 2” OR “SARS CoV 2” OR SARSCoV OR SARS-CoV OR SARS2 OR “Wuhan coronavirus” OR “Wuhan seafood market pneumonia virus” |
| #4 | #1 or #2 |
| #5 | #4 and #3 |
| Filters applied: | Clinical study, Clinical Trial, Comparative Study, Pragmatic Clinical Trial, Randomized Controlled Trial, From 2019/12/1 to present, Humans |

**Supplementary Table S3. Variables extracted from included studies**

| Study details | Author, year of publication, period of study, setting (outpatient, hospitalised non-ICU, ICU), geographical sites, number of enrolled participants in each arm and attrition details (number taken through to analysis), study question, inclusion, exclusion criteria, follow up period/timing of outcome(s) assessment, Trial registration status |
| --- | --- |
| Participant demographics | Overall: (1) mean/median age, (2) proportion female, (3) mean/median number of days from symptom onset, (4) hospitalisation and covid-19 confirmation to randomisation, (5) proportion with baseline D-dimer concentration unknown, low (<2 ULN as per local laboratory range), or high (≥ 2 times ULN), mean/median D-dimer, (6) proportion of participants at baseline using concomitant antiviral drugs, antiplatelet, corticosteroid drugs, other immune modulators (eg, IL-6 antagonists) (7) mean/median BMI OR proportion with recorded elevated BMI (≥25kg/m^2^) (8) median/mean number of documented comorbidities OR proportion with documented comorbidities (overall and individually: hypertension, diabetes, hyperlipidaemia, coronary artery disease, obstructive airway disease, heart failure, previous ischaemic cerebrovascular accidents, previous haemorrhagic stroke or venous thromboembolism), (9) baseline disease severity (median APACHE II/SOFA score, proportion requiring organ support [as defined above], oxygen requirements (median/mean PF ratio, WHO ordinal scale, sO2 < 94%) at time of randomisation/recruitment) |
| Intervention and comparator | Intervention and comparator anticoagulant (agent, dose, route, duration) |
| Efficacy outcomes^a^ | At 30 days (or other short-term follow up period, trial defined), proportion^b^ of participants in each intervention and comparator arm with:  (1) any venous thrombosis (2) any arterial thrombosis (3) in-hospital/death (4) any thrombosis (5) confirmed deep venous thrombosis (6) confirmed pulmonary embolism (7) confirmed ischaemic cerebrovascular accident (8) confirmed myocardial infarction (9) confirmed mesenteric ischaemia (10) confirmed acute limb ischaemia (11) composite outcome (death or mortality) (12) mean/median duration of supplemental oxygen (13) mean/median duration of hospitalisation (14) mean/median duration of ICU admission (15) proportion requiring new organ support from baseline (overall and separately) and mean/median number of days requiring organ support: high flow nasal cannula, non-invasive mechanical ventilation, mechanical ventilation, extracorporeal membrane oxygenation, inotrope or vasopressor support, renal replacement therapy) |
| Safety outcomes^a^ | At 30 days (or other short-term follow up period, trial defined), proportion† of participants in each intervention and comparator arm with:  (1) Major (2) non-major but clinically significant bleeding events, (3) any bleeding event |
| Assessment of bias | See details in main text |

APACHE II – Acute Physiology and Chronic Health Evaluation score II, BMI – body mass index, IL-6 – interleukin-6, PF – Partial pressure of arterial oxygen to inspired oxygen ratio, SOFA – Sequential Organ Failure Assessment, S02 – saturation of oxygen, ULN – upper limit of normal, WHO – World Health Organisation. ^a^Employing trial-specified definitions for outcomes ^b^Denominator for all proportions: as per intention-to-treat analysis; all randomized participants who received at least one dose of assigned treatment.

**Supplementary Table S4. Included studies: intervention, comparator, enrolment period and setting**

| Study (follow up period) | Sample (ITT analysis) | Intervention | Comparator | Enrolment period | Setting | Geographic site |
| --- | --- | --- | --- | --- | --- | --- |
| INSPIRATION (30 days)[1] | 562 | Intermediate-dose enoxaparin | Standard low-dose enoxaparin prophylaxis | Jul 2020 - Nov 2020 | ICU | 10 academic centres in Iran |
| REMAP-CAP, ACTIV-4a and ATTACC (non-critically ill) (21 days)[2] | 2219 | Therapeutic  enoxaparin or UFH | Usual care thromboprophylaxis (low dose or intermediate dose enoxaparin/UFH) | Apr 2020 - Jan 2021 | Hospitalised, non-ICU | 121 sites in 9 Countries † |
| REMAP-CAP, ACTIV-4a and ATTACC (critically ill) (21 days)[3] | 1098 | Therapeutic  enoxaparin or UFH | Usual care thromboprophylaxis (low dose or intermediate dose enoxaparin/UFH) | Apr 2020 - Jan 2021 | ICU-level support | 121 sites in 9 countries ^a^ |
| RAPID (28 days)[4] | 465 | Therapeutic LMWH or UFH | Standard low-dose, prophylactic LMWH or UFH | May 2020 – Apr 2021 | Hospitalised, non-ICU with elevated D- dimer (moderately ill) | 28 sites in 6 countries^b^ |
| HEP-COVID (30 days)[5] | 253 | Therapeutic enoxaparin | Enoxaparin/UFH/Dalteparin - prophylactic or intermediate-dose | May 2020 – Apr 2021 | Hospitalised, requiring oxygen, with elevated D-dimer or coagulopathy (33% in ICU) | 12 centres, USA |
| ACTIV-4B (45 days)[6] | 278 | Apixaban (therapeutic) | Apixaban (prophylactic) | Sep 2020 - Jun 2021 | Outpatient at time of diagnosis and treatment | 52 centres, USA |
| ACTION (30 days)[7] | 614 | Therapeutic rivaroxaban  (stable patients) or enoxaparin/UFH followed by rivaroxaban (unstable patients) | Standard low-dose prophylactic enoxaparin or UFH | Jun 2020 - Feb 2021 | Hospitalized with elevated D-dimer levels (6% requiring ICU admission). | Brazil |
| Perepu et al. (30 days)[8] | 173 | Intermediate-dose enoxaparin | Standard low-dose prophylactic enoxaparin | Apr 2020 - Jan 2021 | ICU or with lab-confirmed coagulopathy (ISTH Overt DIC score ≥ 3) | 3 centres, USA |
| HESACOVID (28 days)[9] | 20 | Therapeutic enoxaparin | Standard-dose, prophylactic enoxaparin (50%) or UFH (50%) | Apr 2020 - Jul 2020 | ICU | Brazil |
| BEMICOP  (30 days)[10] | 65 | Therapeutic bemiparin | Standard low-dose bemiparin prophylaxis | Oct 2020 - May 2021 | Hospitalised, non-ICU with elevated D dimer | 5 hospitals in Spain |
| Oliynyk et al.(28 days)[11] | 126 | Therapeutic LMWH or UFH | Standard low-dose prophylactic enoxaparin | Jul 2020 - Mar 2021 | ICU; elevated D-dimer, not ventilated at baseline | Ukraine (single site) |

DIC – diffuse intravascular coagulopathy, ICU – intensive care unit, ITT – intention-to-treat, LMWH – low molecular weight heparin, UFH – unfractionated heparin, USA – United States of America. ^a^United States of America, Canada, United Kingdom, Brazil, Mexico, Nepal, Australia, The Netherlands, Spain. ^b^Brazil, Canada, Ireland, Saudi Arabia, United Arab Emirates, United States of America.

**Supplementary Table S5. Included studies: baseline participant characteristics/comorbidities**

| Study (follow up period) | Average age (years) | Female, n (%) | Average BMI (kg/m^2^) | HPT, n (%) | DM, n (%) | COPD/CLD, n (%) | HF, n (%) | Current/previous smoker, n (%) |
| --- | --- | --- | --- | --- | --- | --- | --- | --- |
| INSPIRATION (30 days)[1] | Intermediate-dose: 62 Prophylactic: 61 | 237 (42) | 27 | 131 (23) | 155 (28) | 39 (7) | 13 (2) | NR |
| REMAP-CAP, ACTIV-4a and ATTACC (non-critically ill) (21 days)[2] | Therapeutic: 59 Prophylactic: 59 | 921 (42) | 30 | 993 (45) | 663 (30) | 461 (21) | 244 (11) | NR |
| REMAP-CAP, ACTIV-4a and ATTACC (critically ill) (21 days)[3] | Therapeutic: 60  Prophylactic: 62 | 331 (30) | 30 | NR | 362 (33) | 258 (23) | 89 (8) | NR |
| RAPID (28 days)[4] | 60 | 201 (43) | NR | 225 (48) | 160 (34) | 63 (14) | 15 (3) | 9 (2) |
| HEP-COVID (30 days)[5] | Therapeutic: 66 Prophylactic: 68 | 117 (46) | Therapeutic: 31 Prophylactic dose: 30 | 151 (60) | 94 (37) | 17 (7) | 2 (1) | NR |
| ACTIV-4B (45 days)[6] | Median 52 – 55yrs across 4 arms | 197 (71) | Median range 30 - 31 | 177 (64) | 91 (33) | NR | NR | 92 (33) |
| ACTION (30 days)[7] | Therapeutic: 57  Prophylactic: 57 | 247 (40) | 30 | 302 (49) | 150 (24) | 29 (5) | 13 (2) | 119 (19) |
| Perepu et al. (30 days)[8] | 64 | 76 (44) | 31 | 104 (60) | 64 (37) | 39 (23) | 54 (31) | 73 (42) |
| HESACOVID (28 days)[9] | Therapeutic: 55  Prophylactic: 58 | 4 (20) | Therapeutic: 33  Prophylactic dose: 34 | 7 (35) | 7 (35) | 0 (0) | 2 (10) | NR |
| BEMICOP  (30 days)[10] | Therapeutic: 32  Prophylactic: 33 | 24 (37) | 26 | 22 (34) | 5 (8) | 11 (17) | 4 (6) | 26 (40) |
| Oliynyk et al.(28 days)[11] | Therapeutic LMWH: 70  Prophylactic LMWH: 71  Therapeutic UFH: 71 | 50 (40) | NR | NR | NR | NR | NR | NR |

BMI – body mass index, CLD – chronic lung disease, COPD – chronic obstructive lung disease, DM – diabetes mellitus, HPT – hypertension, HF – heart failure, LMWH – low molecular weight heparin, NR – not reported, UFH – unfractionated heparin

**Supplementary Table S6. Included studies: baseline participant characteristics/disease severity**

| Study (follow up period) | Average number of days from symptom onset to hospitalisation (days) | Average number of days from symptom onset to enrolment (days) | Average baseline APACHEII/ SOFA score | Organ support at baseline | Antiviral use at baseline, n (%) | Antiplatelet use at baseline, n (%) | Corticosteroid use at baseline, n (%) |
| --- | --- | --- | --- | --- | --- | --- | --- |
| INSPIRATION (30 days)[1] | 7 | 7 | APACHEII 8 | Vasopressor support within 72 hours of enrolment:127/562 Fi02 >50% at the time of randomization: 234/562 | 443 (79) | 172 (31) | 524 (93) |
| REMAP-CAP, ACTIV-4a and ATTACC (non-critically ill) (21 days)[2] | NR | NR | NR | Any organ support: 1740/2231 | 811 (37) | 259 (12) | 894 (40) |
| REMAP-CAP, ACTIV-4a and ATTACC (critically ill) (21 days)[3] | NR | NR | APACHEII 14 | NR | 346 (32) | 75 (7) | 884 (81) |
| RAPID (28 days)[4] | Therapeutic: 1.5 Prophylactic: 1.4 | NR | NR | NR | NR | 53 (11) | 323 (69) |
| HEP-COVID (30 days)[5] | NR | NR | NR | NR | 178 (70) | 64 (25) | 204 (81) |
| ACTIV-4B (45 days)[6] | NR | NR | NR | Any organ support: 0/493 | NR | 0 | NR |
| ACTION (30 days)[7] | Therapeutic: 8 Prophylactic: 7 | 10 | NR | Oxygen support: 460/615  Mechanically ventilated: 39/615 | 10 (2) | 48 (8) | 510 (83) |
| Perepu et al. (30 days)[8] | NR | NR | NR | In ICU at baseline: 107/173  Mechanical ventilation: 40/173 | 105 (61) | NR | 130 (75) |
| HESACOVID (28 days)[9] | Therapeutic: 7 Prophylactic: 6 | NR | SOFA 10 | Mechanical ventilation: 20/20 | 0 | 0 | 14 (70) |
| BEMICOP  (30 days)[10] | NR | 8 | NR | Oxygen support: 38/65 | 9 (14) | 0 | 62 (95) |
| Oliynyk et al.(28 days)[11] | NR | NR | NR | Oxygen support: 126/126  (Inclusion criteria PaO_2_ < 60mmHg, but not mechanically ventilated) | NR | NR | NR |

APACHEII - Acute Physiology and Chronic Health Evaluation score II, Fi02 – fraction of oxygen in inspired air, NR – not reported, PaO_2_ – partial pressure of oxygen in arterial blood, SOFA – Sequential Organ Failure Assessment

**Supplementary Table S7. Assessment of bias in individual studies**

| **Study (follow up period)** | **Risk of bias arising from randomization process** | **Risk of bias due to deviations from the intended interventions (effect of assignment to invention)** | **Risk of bias due to deviations from the intended interventions (effect of adhering to intervention)** | **Risk of bias due to missing outcome data** | **Risk of bias in measurement of outcome** | **Risk of bias in selection of reported results** | **Overall risk** |
| --- | --- | --- | --- | --- | --- | --- | --- |
| **INSPIRATION (30 days)[1]** |  |  |  |  |  |  |  |
| **REMAP-CAP, ACTIV-4a and ATTACC (non-critically ill) (21 days)[2]** |  |  |  |  |  |  |  |
| **REMAP-CAP, ACTIV-4a and ATTACC (critically ill) (21 days)[3]** |  |  |  |  |  |  |  |
| **RAPID (28 days)[4]** |  |  |  |  |  |  |  |
| **HEP-COVID (30 days)[5]** |  |  |  |  |  |  |  |
| **ACTIV-4B (45 days)[6]** |  |  |  |  |  |  |  |
| **ACTION (30 days)[7]** |  |  |  |  |  |  |  |
| **Perepu et al. (30 days)[8]** |  |  |  |  |  |  |  |
| **HESACOVID (28 days)[9]** |  |  |  |  |  |  |  |
| **BEMICOP**  **(30 days)[10]** |  |  |  |  |  |  |  |
| **Oliynyk et al.(28 days)[11]** |  |  |  |  |  |  |  |

**Low	Some concerns	High risk**

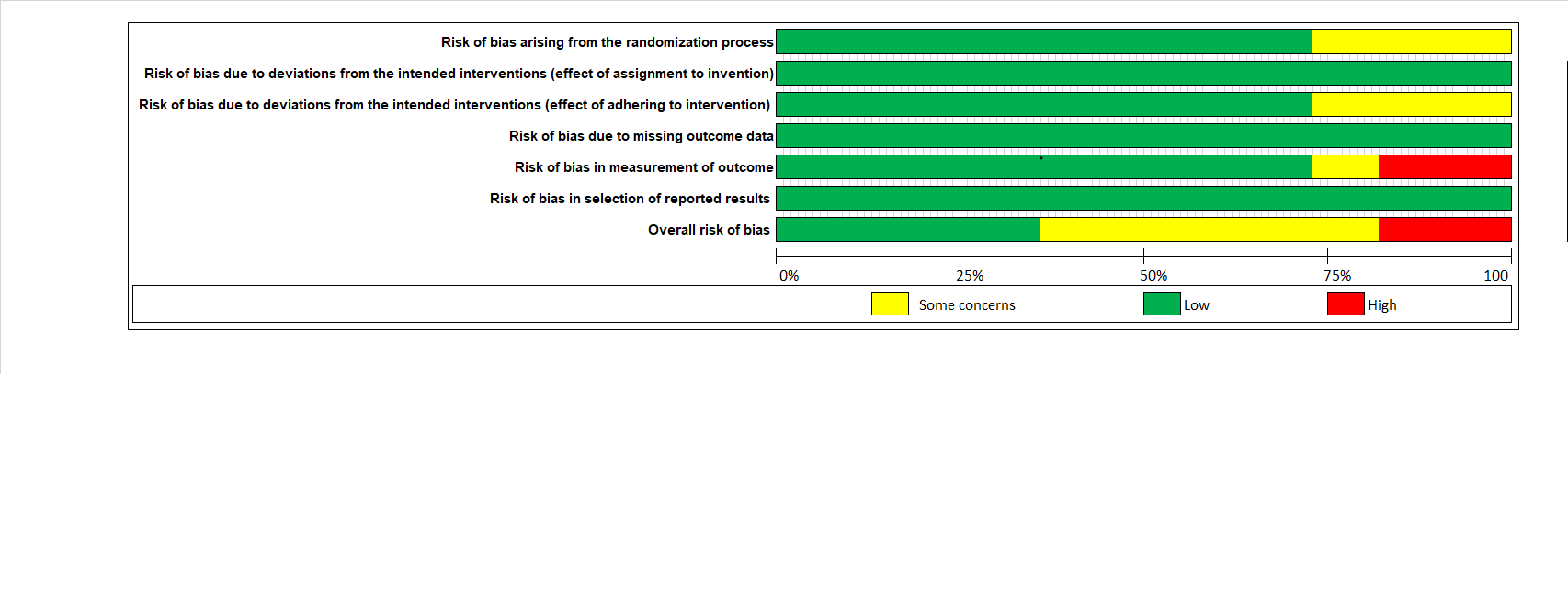

**Supplementary Figure S1. Risk of bias graph: review authors' judgements about each risk of bias item presented as percentages across all included studies.**

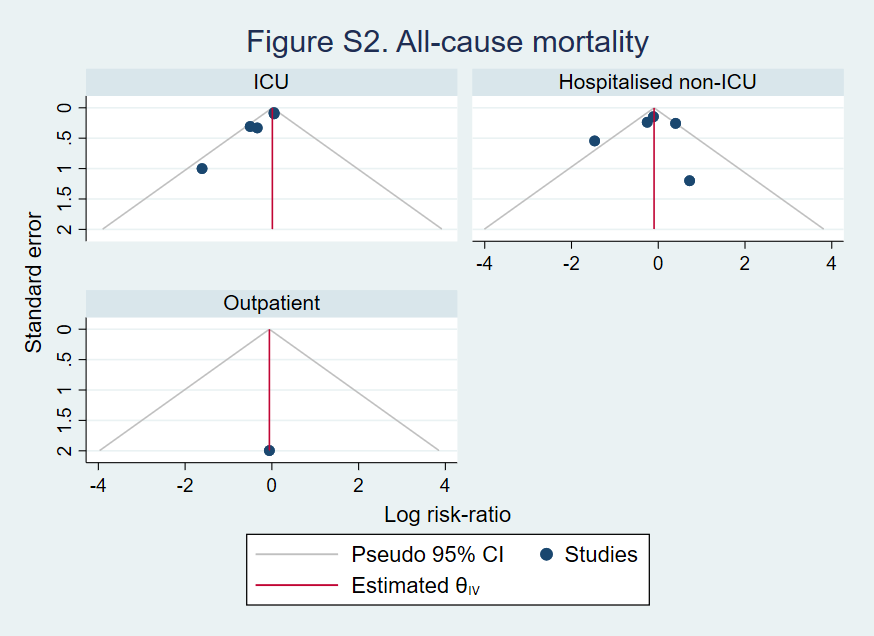

**Supplementary Figure S2. Funnel plot for mortality in groups receiving intensified versus prophylactic anticoagulation, stratified by clinical setting (ICU, hospitalised non-ICU and outpatient) (n = 11 studies)**

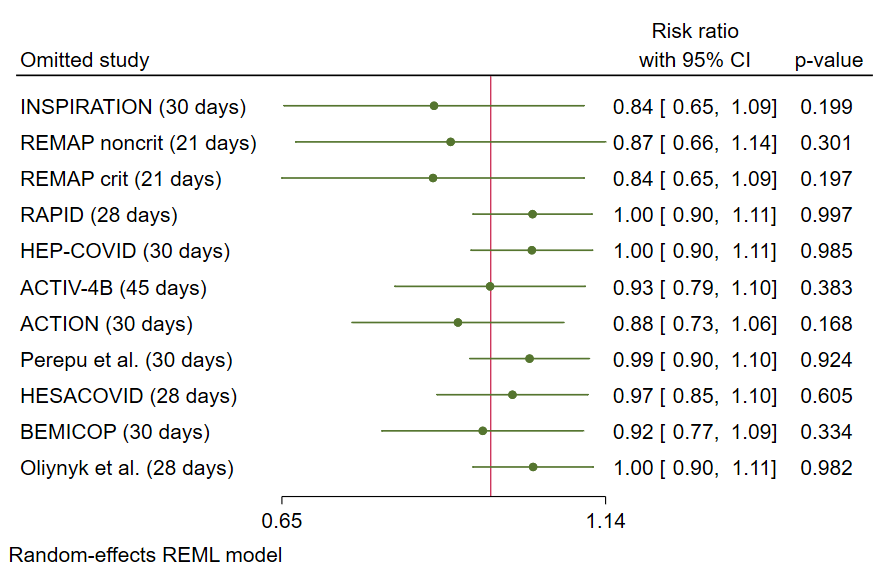

**Supplementary Figure S3. Leave-one-out forest plot, demonstrating overall effect size computed from meta-analysis with each individual trial excluded (n = 11 studies)**

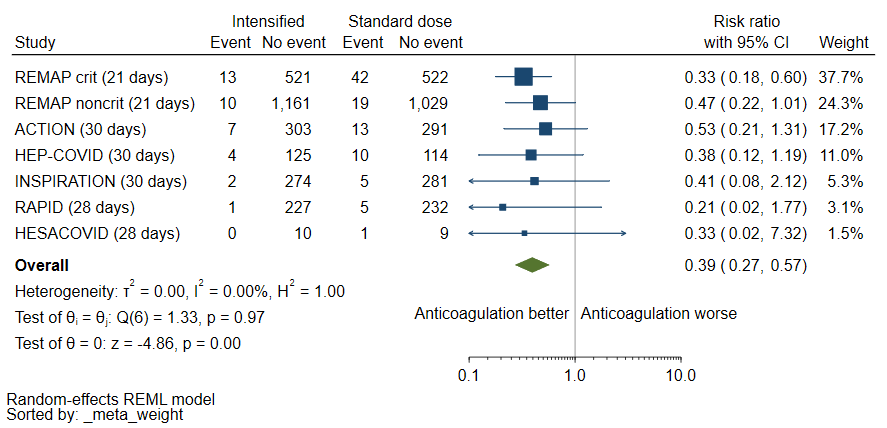

**Supplementary Figure S4. Forest plot of risk ratio for pulmonary embolism (PE) in groups receiving intensified versus prophylactic anticoagulation**

**(n = 7 studies).** There were no thrombotic events in the single outpatient trial[6] and the pulmonary embolic events were not captured as outcomes in the remaining trials[8,10,11] – these trials are excluded from the forest plot above.

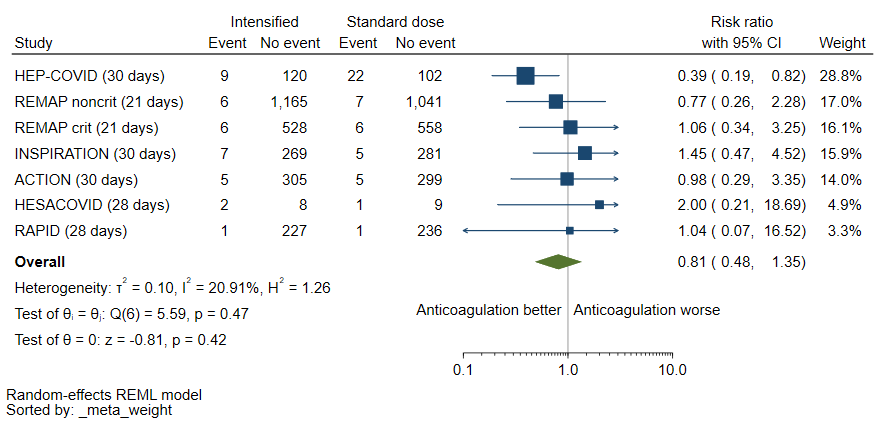

**Supplementary Figure S5. Forest plot of risk ratio for deep venous thrombosis in groups receiving intensified versus prophylactic anticoagulation**

**(n = 7 studies).** There were no thrombotic events in the single outpatient trial[6] and DVT events were not captured as outcomes in the remaining trials[8,10,11] – these trials are excluded from the forest plot above.

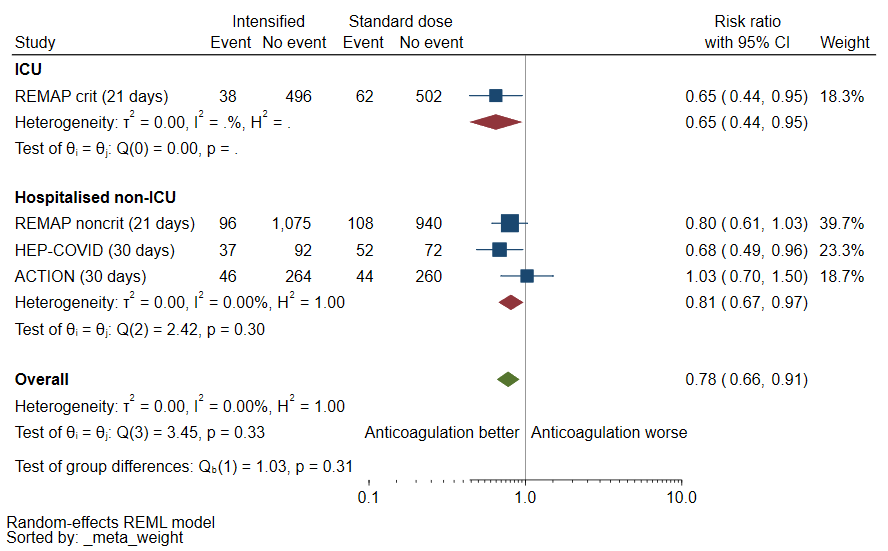

**Supplementary Figure S6. Forest plot of risk ratio for composite outcome (any thrombosis or death) in groups receiving intensified versus prophylactic anticoagulation, stratified by clinical setting (ICU versus hospitalised non-ICU) (n = 4 studies).** Only 4 studies reported on this composite outcome,[2,3,5,7] all comparing therapeutic anticoagulation to low or intermediate-dose prophylactic anticoagulation.

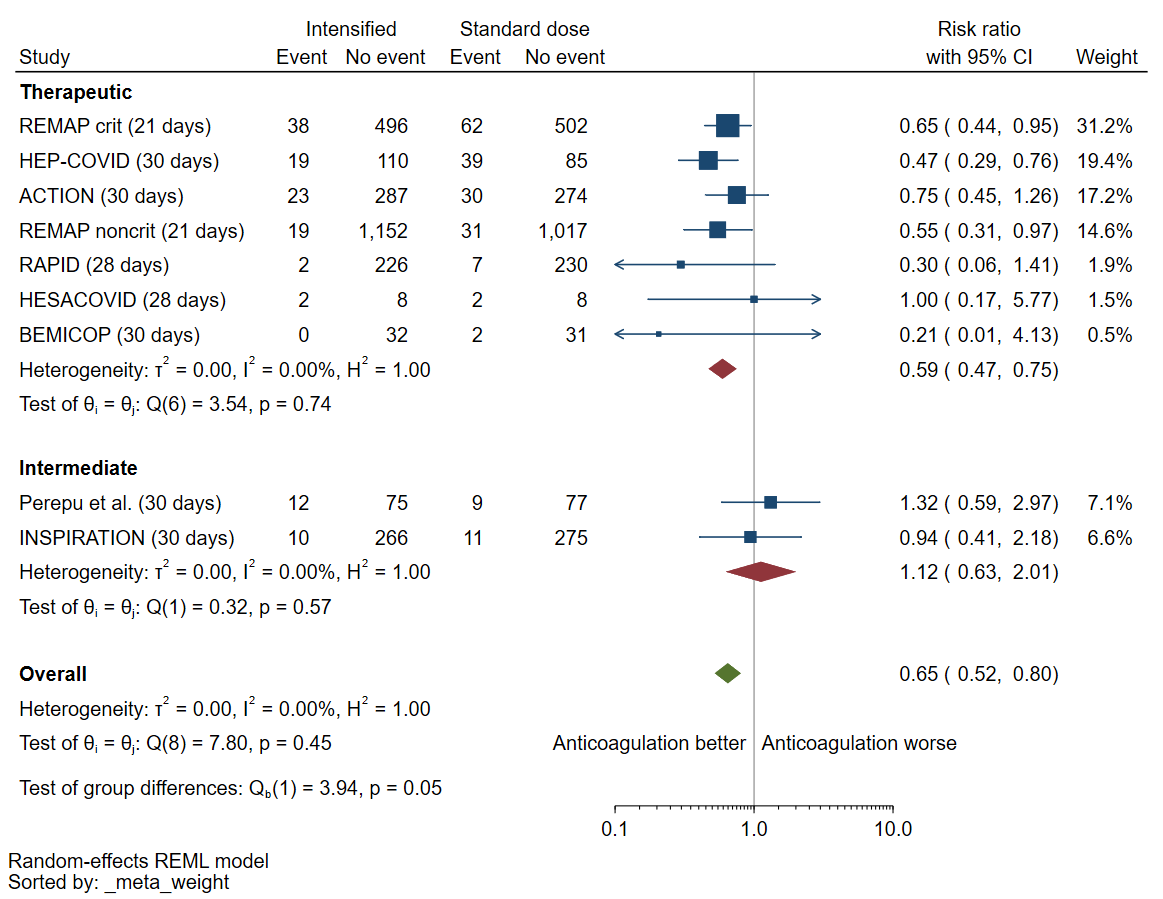

**Supplementary Figure S7. Forest plot of risk ratio for any thrombosis in groups receiving intensified versus prophylactic anticoagulation, stratified by dose of intensified anticoagulation (therapeutic versus intermediate) (n = 9 studies).** There were no thrombotic events in the single outpatient trial[6] and thrombotic events were not captured as an outcome in another trial [11] – these trials are excluded from the forest plot above.

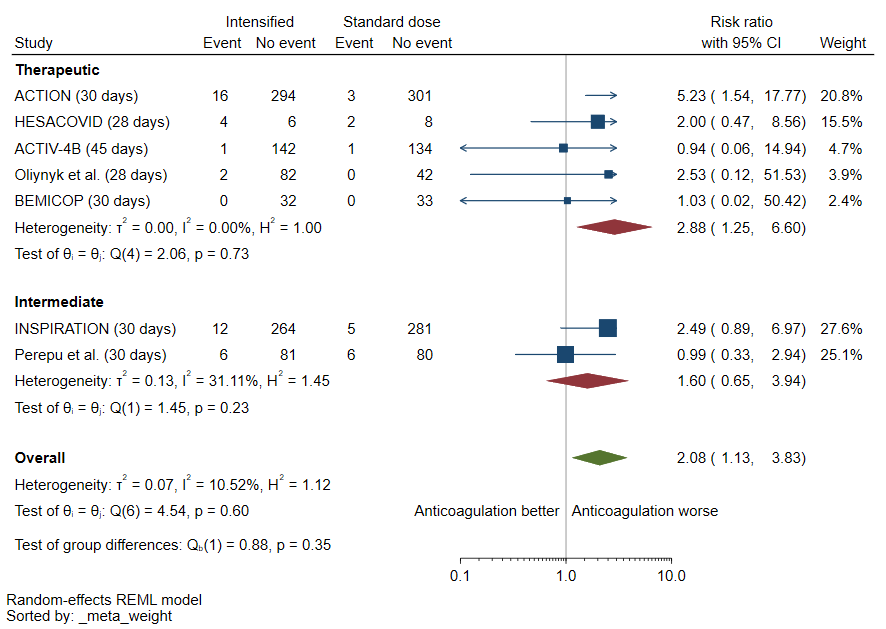

**Supplementary Figure S8. Forest plot of risk ratio for clinically relevant non-major bleed in groups receiving intensified versus prophylactic anticoagulation, stratified by dose of intensified anticoagulation (therapeutic versus intermediate) (n = 7 studies).** This outcome was not reported by 4 trials[2–5] excluded from forest plot above.

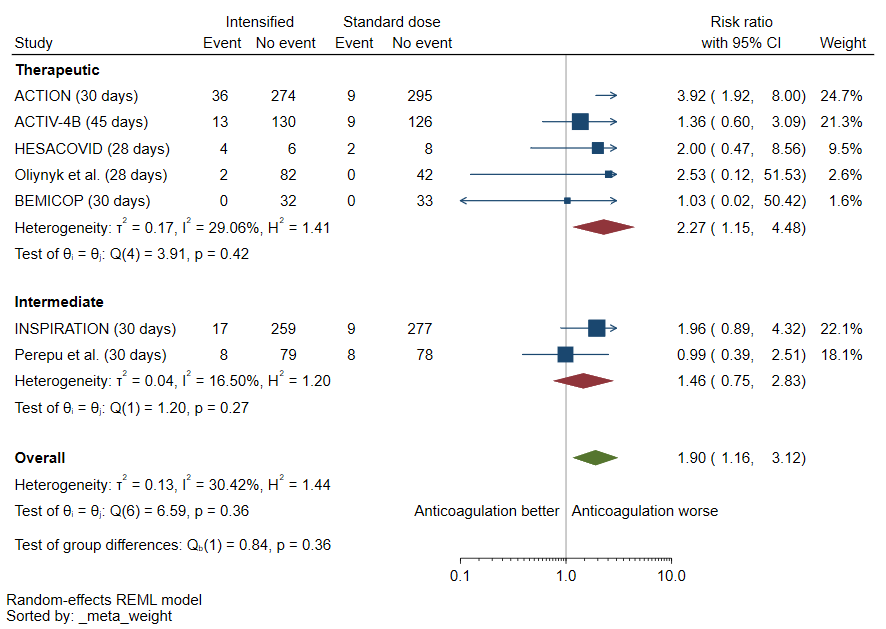

**Supplementary Figure S9. Forest plot of risk ratio for any bleed in groups receiving intensified versus prophylactic anticoagulation, stratified by dose of intensified anticoagulation (therapeutic versus intermediate) (n = 7 studies).** This outcome was not reported by 4 trials[2–5] excluded from forest plot above.

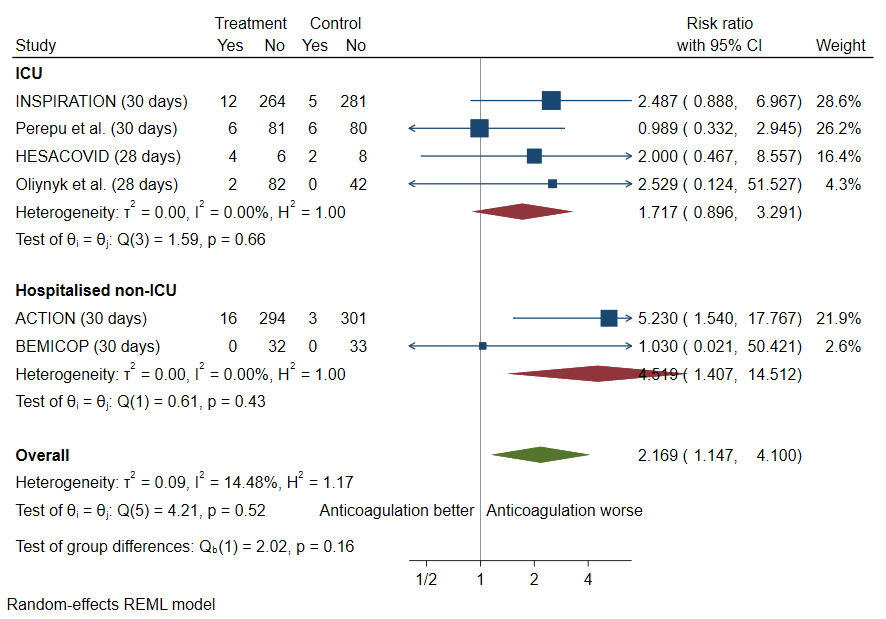

**Supplementary Figure S10. Forest plot of risk ratio for clinically relevant non-major bleed in groups receiving intensified versus prophylactic anticoagulation, stratified by clinical setting (ICU versus hospitalised non-ICU) (n = 6 studies).** This outcome was not reported by 4 in-hospital trials [2–5] excluded from the forest plot above.

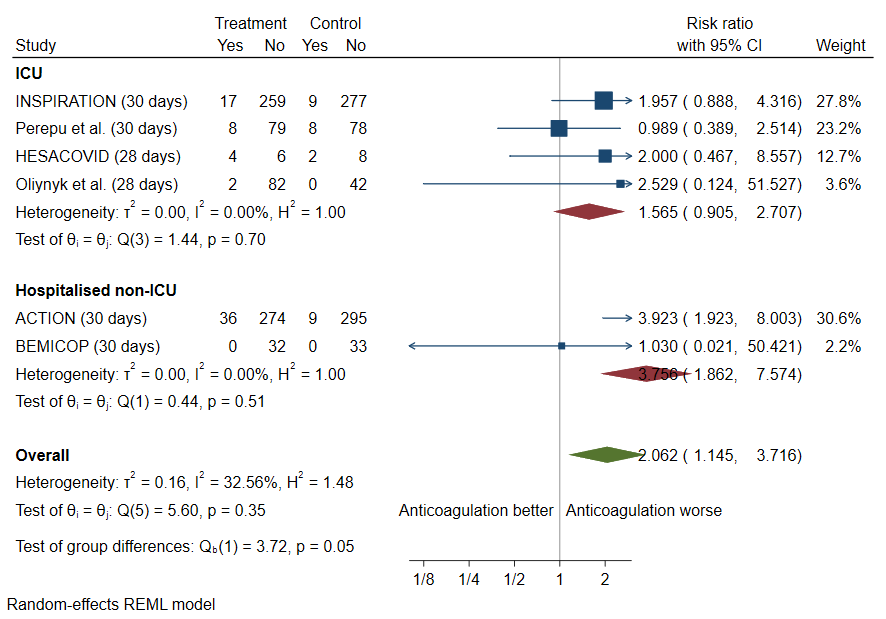

**Supplementary Figure S11. Forest plot of risk ratio for any bleed in groups receiving intensified versus prophylactic anticoagulation, stratified by clinical setting (ICU versus hospitalised non-ICU)(n = 6 studies).** This outcome was not reported by 4 in-hospital trials [2–5] excluded from the forest plot above.

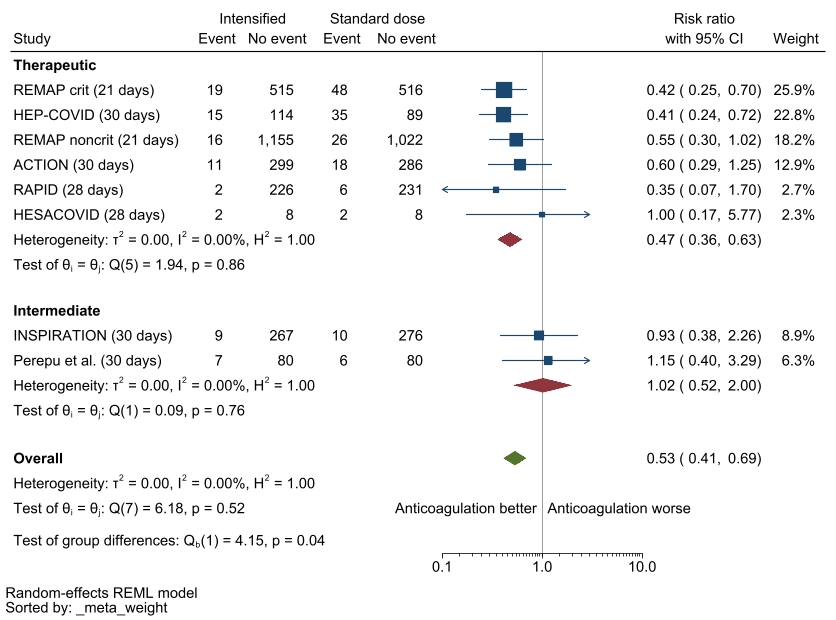

**Supplementary Figure S12. Forest plot of risk ratio for venous thrombosis in groups receiving intensified versus prophylactic anticoagulation, stratified by dose of intensified anticoagulation (therapeutic versus intermediate) (n = 8 studies).** There were no reported thrombotic events in the single outpatient trial[6] and venous thromboembolic events were captured as outcomes in two trials[10,11] – these trials are excluded from the forest plot above.
